# CONTRAST MEDIA VOLUME CONTROL FOR LIMITING ACUTE KIDNEY IN ACUTE CORONARY SYNDROME: THE REMEDIAL IV RANDOMIZED CLINICAL TRIAL

**DOI:** 10.1101/2023.09.28.23296315

**Authors:** Carlo Briguori, Cristina Quintavalle, Enrica Mariano, Alessandro D’Agostino, Mario Scarpelli, Amelia Focaccio, Giuseppe Biondi Zoccai, Salvatore Evola, Giovanni Esposito, Giuseppe Massimo Sangiorgi, Gerolama Condorelli

## Abstract

**BACKGROUND:** Acute kidney injury (AKI) is a common complication in patients suffering from acute coronary syndromes (ACS) and treated by percutaneous coronary intervention (PCI). Contrast media (CM) volume minimization has been advocated to prevent AKI. The DyeVert™ system (Osprey Medical Inc., Minnetonka, MN, USA) is a device designed to reduce CM volume during coronary procedures, while maintaining fluoroscopic image quality.

**METHODS:** In this is study a randomized, single-blind, investigator-driven clinical trial conducted in 4 italian interventional cardiology centers from February 4, 2020 to September 13, 2022, 550 ACS participants were randomly assigned in 1:1 ratio to 1) *Control group* (n = 274), in which a conventional manual or automatic injection syringe was used, and 2) *Contrast Volume Reduction (CVR) group* (n = 276), in which CM injection was handled by the DyeVert^TM^ system. The primary endpoints were 1) CM volume, and 2) the rate of AKI, defined as a serum creatinine (sCr) increase ≥0.3 mg/dL within 48 hours after CM exposure.

**RESULTS:** There were 412/550 (74.5%) participants with ST-elevation myocardial infarction (201/274 [73.3%] in the *Control group* and 211/276 [76.4%] in the *CVR group*). Mean glomerular filtration rate was 84±32 mL/min/1.73 m^2^ in the *Control group* and in 85±34 mL/min/1.73 m^2^ in the *CVR group* (p = 0.78). CM volume was higher in the *Control group* (160 ± 23 mL versus 95 ± 30 mL; p < 0.001). Seven participants (6 in the *Control group* and one in the *CVR group*) did not have post-procedural sCr values. AKI occurred in 65/268 (24.3%) participants in the *Control group* and in 44/275 (16%) participants in the *CVR group* (RR = 0.66; 95% confidence interval 0.47-0.93; p = 0.018).

**CONCLUSIONS:** CM volume reduction obtained by the DyeVert^TM^ system is effective to prevent AKI in ACS patients undergoing invasive procedure.

**Trial Registration:** The study is registered with www.clinicaltrial.gov (NCT04714736)

**Clinical Perspective:** *What Is New?:*

- Contrast media volume reduction obtained by the DyeVert^TM^ system is effective to prevent acute kidney injury in acute coronary syndrome patients undergoing invasive procedure.

*What Are the Clinical Implications?:*

- Contrast media volume minimization is of outmost important in the attempt to prevent acute kidney injury. The DyeVert™ system is an “operator-independent” tool contributing to the contrast media-sparing approach.

## INTRODUCTION

Acute kidney injury (AKI) may occur in patients suffering from acute coronary syndromes (ACS) and treated by percutaneous coronary intervention (PCI) ^1–3^.. Although the pathogenesis of AKI in ACS patients undergoing PCI is multifactorial ^4^, the role of iodinated contrast media (CM) has been well established^5^. Volume expansion represents the cornerstone in contrast-associated AKI (CA-AKI) prevention^6^. However, all the recommended volume expansion regimens have limited applicability in ST-Elevation Myocardial Infarction (STEMI) and high-risk Non-ST-Elevation Myocardial Infarction (NSTEMI) patients transferred to percutaneous coronary intervention (PCI)-capable centres for emergency invasive treatment. Therefore, in this scenario is of outmost importance CM volume minimization in the attempt to prevent CA-AKI. The DyeVert™ system (Osprey Medical Inc., Minnetonka, MN, USA) is a device designed to reduce CM volume during coronary procedures, while maintaining fluoroscopic image quality^7^.

The aim of the REnal Insufficiency Following Contrast MEDIA Administration triaL IV (REMEDIAL IV) is to test whether the use of the DyeVert system is effective in reducing AKI rate in ACS patients undergoing urgent invasive approach.

## METHODS

### Patient population

This is a multicenter, randomized, investigator-driven, clinical trial, whose design has been previously reported ^8^. All patients with STEMI and high-risk NSTEMI requiring urgent/immediate invasive approach were screened for inclusion/exclusion criteria (Tables S1-S3) ^9–11^. Cardiogenic shock was defined according to the Society for Cardiovascular Angiography and Interventions (SCAI) classification ^12^ (Table S4). All participants or their legally authorized representatives provided written informed consent. The trial was registered with www.clinicaltrial.gov (NCT04714736), was conducted at 4 Italian interventional cardiology centers (Table S5), according to the principles of the Declaration of Helsinki ^13^ and Good Clinical Practice ^14^ and has been approved by the local Ethic Committees.

Participants were randomly assigned to:

#### Contrast volume reduction (CVR) group

CM injection was handled by the DyeVert^TM^ system, a device designed to reduce CM volume during coronary procedures, while maintaining fluoroscopic image quality ^7^. During an injection, the DyeVert^TM^ system diverts a portion of the injected CM through a secondary fluid pathway controlled by a pressure-compensating diversion valve. This allows a decrease in over-injection of CM and less aortic reflux. The valve is constructed in a way that the diversion pathway resistance automatically increases with higher injection pressures and decreases with lower injection pressures proportionally decreasing or increasing CM delivered to the patient, respectively. The diverted CM is temporarily stored in the reservoir (Supplementary Data). The associated Contrast Monitoring System displays CM volume injected (in milliliters, mL), split in attempted, delivered and saved (the last reported both as absolute value and as percentage versus the total). Previous studies reported that the DyeVert^TM^ system allows an up to 40% reduction in CM volume ^15, 16^.

#### Control group

CM injection in this group was carried out by a conventional manual injection syringe or automatic injection device (ACIST Medical System, Eden Prairie, Minnesota, USA).

### Volume expansion regimen

Normal saline (3 mL/kg/h) was initiated as soon as the participants arrived in the catheterization laboratory. Intraprocedural volume expansion rate was adjusted according to the left ventricular end-diastolic pressure (LVEDP), which was estimated at the beginning of the procedure: 5 mL/kg/h for LVEDP ≤12 mmHg; 3 mL/kg/h for LVEDP 13-18 mmHg; and 1.5 mL/kg/h for LVEDP >18 mmHg ^17^. When deemed clinically contraindicated (pulmonary congestion, acute heart failure [Table S3]) volume expansion was not started at all. Volume expansion continued during the procedure and for at least 6 hours post-procedure. Total hydration >960 mL was consider the optimal cutoff volume to prevent CA-AKI ^18^.

### Biomarkers of kidney function

Serum creatinine (sCr), cystatin C, blood urea nitrogen, sodium and potassium were measured at baseline (that is, as soon as the participant arrived in the emergency room and/or in the catheterization laboratory room before intravenous volume expansion initiation and CM injection), and every day during the hospital stay; additional measurements were performed in all cases of deterioration of baseline renal function. Estimated glomerular filtration rate (GFR) was calculated by applying Chronic Kidney Disease Epidemiology Collaboration (CKD-EPI) equation ^19^. The risk scores for predicting CA-AKI were evaluated according to Mehran’s score ^20^ and Gurm’s score ^21^ (Tables S6-S7).

### Iodinated contrast media

Iobitridol (Xenetix®350, 350 mg iodine/mL, Guerbet, France) a non-ionic, low-osmolality (915 mOsm per kilogram of water) CM was used in all instances. Strategies for limiting CM volume were implemented^22^ . The administration of a CM volume >3X GFR is suggestive of increased risk of CA-AKI ^23^. Number of injections (including ‘‘tests’’ and ‘‘puffs’’) per patient was analyzed. Radiation exposure per patient was measured as 1) dose rate (the amount of radiation delivered per unit time), expressed as grays (Gy) and 2) dose area product (DAP; Gy/cm^2^).

### Study endpoints

The primary endpoints are 2) the CM volume, and 2) the rate of CA-AKI, defined, according to the Kidney Disease: Improving Global Outcomes (KDIGO) criteria, an increase in the sCr concentration ≥0.3 mg/dL within 48 hours ^24^. Secondary end-points included: 1) an increase in the sCr concentration ≥25% and/or ≥0.5 mg/dL within 72 hours after CM exposure; 2) the severity of AKI assessed according to the KDIGO criteria^24^; 3) changes in the serum cystatin C concentration at 24 and 48 hours after CM exposure; 4) the rate of acute renal failure requiring renal replacement therapy (RRT, defined as a decrease in renal function necessitating acute hemodialysis, ultrafiltration or peritoneal dialysis within the first 5 days post-intervention); 5) the length of in-hospital stay, calculated as the sum of the number of days since admission until discharge from the hospital; and 6) the rate of in-hospital, 1, and 6-month major adverse events, including death, non-fatal myocardial infarction, RRT, sustained kidney injury and major bleeding. Major bleeding were defined according to the BARC criteria ^25^. Sustained kidney injury was defined as a persistent ≥25% GFR reduction compared to baseline at the last available value during the follow up ^26^. All events were adjudicated by a Clinical Events Committee (CEC), who was blinded to treatment assignment.

### Statistical analysis

Treatment allocation to the 2 groups was determined by randomization in a 1:1 ratio. To ensure that almost equal number of participants receive one of the 2 treatments a randomization blocks of 4 was used. An independent statistician generated the randomization list with permuted blocks, and the block size was not disclosed to the investigators enrolling the participants (Random Allocation Software 1.0). Participants were randomly assigned without stratification by STEMI vs NSTEMI. According to published data, the expected CA-AKI rate in the *Control group* is 19% ^1–3, 16, 18^. A sample size in each group of 261 patients (a total of at least 522 randomized participants) was therefore needed to demonstrate a 8.5% difference between groups (that is from 19% in the *Control group* to 10.5% in the *CVR group*), with a two-sided 95% confidence interval (CI) and 80% power (p<0.05); based on the large sample normal approximation extended 0.07 from the observed difference in proportions ^16,28^. Taking into account a dropout rate ≤5%, we recruited 550 participants.. All principal analyses were performed in the intention-to-treat population, defined as all participants randomized, regardless of the treatment actually received.

Continuous variables are given as mean ± 1 standard deviation or median and first and third quartiles (Q1-Q3) and compared using the Student’s t test or Mann-Whitney U test, respectively. Normality assumption was verified graphically (i.e., QQ plot) and was confirmed using the Shapiro-Wilk test. Categorical variables were reported as percentage and were analyzed by either Chi-squared or Fisher’s exact test, as appropriate. We calculated the relative risk and absolute risk difference, and their 95% confidence intervals (CI), for the primary and secondary endpoints. The inverse of the absolute risk difference yielded the number needed to treat (NNT) to prevent one event. A logistic regression was done to assess interaction between CM volume, treatment group and CA-AKI. Variance Inflation Factors analysis was implemented to exclude collinearity. Hosmer-Lemeshow goodness-of-fit test was assessed. To assess the impact the 2 treatments on sCr and cystatin C, we used repeated measures analysis of variance (RM-ANOVA) models, after transforming sCr and cystatin C levels into natural logarithm (to overcome the problem of the non-normal distribution). Probability level <0.05 was considered significant. Statistical analyses were performed using SPSS for Windows, release 20.0 (SPSS Inc., Chicago, Illinois) and Stata 11.2 for Windows (Stata Corp. LP).

## RESULTS

### Patients population

From February 4, 2020 to September 13, 2022, 550 participants were enrolled into the study. The number of participants screened, treated and analyzed are reported according to the CONSORT guidelines (Figure 1). The clinical and biochemical characteristics were well matched between the two groups (Table 1). The proportion of STEMI and high-risk NSTEMI is reported in the Table 1. Mean eGFR was 84±32 mL/min/1.73 m^2^ in the *Control group* and in 85±34 mL/min/1.73 m^2^ in the *CVR group* (p = 0.78). LVEF <40% was observed in 74 (27%) participants in the *Control group* and 74 (26.8%) in the *CVR group* (p = 1.00). Details on PCI are reported in the Table 2. Radial approach was used in the majority of patients. Automatic injection was used in 20 (7.3%) participants in the *Control group* and in 21 (7.6%) in the *CVR group* (p = 1.00).

**Figure 1.**
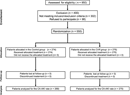
Diagram showing the flow of participants through each stage of the trial according to the Consolidated Standards of Reporting Trials (CONSORT) guidelines. Six patients in the *Control group* and one patients in the *CVR group* were not analyzed for the primary endpoint because they died within 24 (3-22) hours after enrolment and therefore they did not have at least one in-hospital post-procedural serum creatinine value.

**Table 1.** Baseline characteristics and clinical presentation of the participants enrolled into the 2 groups.

|  | **Control group**<br>**(n= 274)** | **CVR group**<br>**(n= 276)** |
| --- | --- | --- |
| **Age (years)** | 65 ± 12 | 64 ± 13 |
| **≥75** | 64 (23.5%) | 64 (23.0%) |
| **Male** | 202 (73.0%) | 220 (80.0%) |
| **Weight (Kg)** | 78±15 | 79±17 |
| **Height (m)** | 1.68±0.2 | 1.69±0.9 |
| **Body-mass Index (Kg/m<sup>2</sup>)** | 27 ± 4 | 27 ± 7 |
| **Blood pressure (mmHg)** |  |  |
| **Systolic** | 127±26 | 125±27 |
| **Diastolic** | 77±14 | 77±17 |
| **Mean** | 110±19 | 109±20 |
| **Diabetes mellitus** | 57 (21.0%) | 58 (21.0%) |
| **Insulin-treated** | 18 (6.5%) | 18 (6.5%) |
| **Peripheral artery disease** | 20 (7%) | 23 (8.0%) |
| **Systemic hypertension** | 165 (60.5%) | 154 (56%) |
| **Previous myocardial infarction** | 24 (8.8%) | 17 (6.1%) |
| **Previous TIA or stroke** | 6 (2.2%) | 13 (4.5%) |
| **Previous percutaneous coronary intervention** | 41 (15.2%) | 41 (15.0%) |
| **Previous coronary artery bypass surgery** | 6 (2.0%) | 8 (1.5%) |
| **Left ventricular ejection fraction (%)** | 45 ± 11 | 45 ± 10 |
| **Type of myocardial infarction** |  |  |
| **ST-elevation myocardial infarction** | 201 (73.5%) | 211 (76.5%) |
| **Non ST-elevation myocardial infarction** | 73 (26.5%) | 65 (23.5%) |
| **Cardiac arrest** | 14 (5.0%) | 13 (4.7%) |
| **Cardiogenic shock** | 18 (6.5%) | 14 (5.0%) |
| **Stage A** | 2 (0.7%) | 2 (0.7%) |
| **Stage B** | 5 (1.8%) | 2 (0.7%) |
| **Stage C** | 11 (4.0%) | 10 (3.6%) |
| **Ongoing medical therapy** |  |  |
| **ACE inhibitor** | 59 (21.5%) | 58 (21.0%) |
| **Angiotensin II receptor inhibitor** | 27 (10%) | 30 (11%) |
| **Diuretics** | 16 (6.0%) | 19 (7.0%) |
| **Beta blockers** | 57 (21.0%) | 55 (20.0%) |
| **Statins** | 123 (45.0%) | 118 (43.0%) |
| **Serum creatinine, mg/dL (median; Q1-Q3)** | 0.96 (0.82-1.19) | 0.99 (0.82-1.18) |
| **eGFR (mL/min/1.73 m<sup>2</sup>)** | 84 ± 32 | 85 ± 34 |
| **<60** | 62 (22.7%) | 54 (19.5%) |
| **Serum cystatin C, mg/dL (median, Q1-Q3)\*** | 1.01 (0.79-1.38) | 1.00 (0.80-1.18) |
| **Serum urea nitrogen, mg/dL** | 44.6±20.9 | 44.5±23.7 |
| **Serum sodium, mEq/L** | 137.8±7.1 | 138.7±3.4 |
| **Serum potassium, mEq/L** | 4.0±0.6 | 4.1±0.7 |
| **Hemoglobin, g/dL** | 14 ± 2 | 14 ± 2 |
| **C reactive protein, median (Q1-Q3)** | 5.0 (1.0-13.0) | 5.0 (1.0-14.5) |
| **Predicted risk for CA-AKI** |  |  |
| **Gurm risk score** | 10 ± 8 | 10 ± 9 |
| ≥7 | 145 (53.0%) | 138 (50.0%) |
| **Mehran risk score** | 9 ± 3 | 9 ± 3 |
| ≥12 | 57 (21.0%) | 68 (25.0%) |

ACE = angiotensin-converting enzyme; TIA = transient ischemic attack. CA-AKI = contrast-associated acute kidney injury; eGFR = estimated glomerular filtration rate;\*Cystatin C was available in 162/274 (59.1%) of participants in the *Control group* and 164/276 (58.6%) participants in the *CVR group*.

**Table 2.** Procedural characteristics of the participants enrolled into the 2 groups.

|  | **Control group**<br>**(n= 274)** | **CVR group**<br>**(n= 276)** |
| --- | --- | --- |
| **Coronary angiography attempted** | 274 (100%) | 276 (100%) |
| **Coronary angiography completed** | 274 (100%) | 276 (100%) |
| **PCI attempted** | 235 (85.7%) | 243 (88.0%) |
| **PCI completed** | 235 (85.7%) | 243 (88.0%) |
| **Radial approach** | 268 (98.5%) | 265 (96.5%) |
| **Symptoms onset to treatment, hour (median, IQR)** | 5.0 (3.2-9.0) | 5.1 (3.2-9.4) |
| **ST-elevation myocardial infarction** | 4.0 (2.1-7.0) | 4.0 (2.2-8,3) |
| **Non ST- elevation myocardial infarction** | 5.5 (3.3-8.2) | 7.1 (3.3-9.5) |
| **Left ventricular end diastolic pressure (mmHg)** | 16 ± 7 | 17 ± 6 |
| **≤12** | 86 (31.5%) | 73 (26.0%) |
| **13-18** | 103 (38.0%) | 96 (35.0%) |
| **>18** | 85 (31.0%) | 107 (39.0%) |
| **Mechanical assist device** | 16 (5.8%) | 14 (5.0%) |
| **Intraaortic balloon pump** | 1 (0.4%) | 1 (0.3%) |
| **Impella CP** | 15 (5.4%) | 13 (4.7%) |
| **Vasopressors** | 29 (10.6%) | 22 (8.0%) |
| **Noradrenaline** | 23 (8.4%) | 11 (4.0%) |
| **Adrenaline** | 6 (2.2%) | 11 (4.0%) |
| **Multivessel disease** | 112 (41.0%) | 115 (41.5%) |
| **Vessel treated** | 290 | 292 |
| **Left main** | 16 (5.5%) | 14 (4.8%) |
| **Left anterior descending artery** | 140 (48.2%) | 144 (49.3%) |
| **Left circumflex artery** | 38 (13.1%) | 39 (13.3%) |
| **Right coronary artery** | 81 (27.9%) | 82 (28.1%) |
| **Others** | 15 (5.1%) | 13 (4.5%) |
| **Number of lesion treated per patient** | 1.31±0.59 | 1.37±0.58 |
| **Total stent length, mm** | 41.8±31.2 | 42.8±27.4 |
| **Multivessel stenting** | 52/235 (22.1%) | 50/243 (20.5%) |
| **Number of stent implanted** | 1.53 ± 0.91 | 1.56 ± 0.95 |
| **0** | 2/235 (0.9%) | 3/243 (1.2%) |
| **1** | 139/235 | 146/243 (60.0%) |
| **2** | (59.1%) | 57/243 (23.5%) |
| **≥3** | 67/235 (28.5%) | 37/243(15.3%) |
|  | 27/235 (11.5%) |  |
| **Radiation exposure** |  |  |
| **Dose rate (Gy)** | 1.53±1.32 | 1.38±1.07 |
| **Dose area product (Gy/cm<sup>2</sup>)** | 118±108 | 112±94 |
| **Fluoroscopic time (minutes)** | 14±11 | 14±10 |

### Volume expansion

Mean 24-hour volume expansion was similar in the 2 groups (1669±517 mL in the *Control group* versus 1691±473 mL *CVR group*; p = 0.61). In particular, volume was >960 mL in 254 (92.5%) participants in the *Control group* and in 257 (93.2%) participants in the *CVR group* (p = 0.74). Peri-procedural intravenous furosemide was administered in 85 (31%) participants in the *Control group* and in 78 (28%) participants in the *CVR group* (p = 0.45). Daily diuresis was similar on the 2 groups both at 24 hours (*Control group* 1577±799 mL versus *CVR group* 1613±794 mL; p = 0.59), at 48 hours (2112±855 mL versus 2061±936 mL; p = 0.52) and at 72 hours (2130±8932 mL versus 2153±958 mL; p = 0.80).

### Primary endpoints

CM volume was higher in the *Control group* than in the *CVR group* (160 ± 23 mL versus 95 ± 30 mL; p < 0.001; Figure 2, panel A). A CM volume >3X GFR was reported in 62 (22.7%) participants in the *Control group* and in 25 (10.7%) in the *CVR group* (p < 0.001). In the *CVR group* the mean absolute and percent CM volume saved was 59.8 ± 37.3 mL and, respectively, 38.1 ± 9.4%. This finding was similar in the subgroup of participants in which automatic injection was used (36.7 ± 8.2%). CM volume saved was higher in patients with multivessel than in single-vessel PCI (75±33 mL vs. 45±37mL; p = 0.025). The DyeVert system was not turned off under any circumstances due to inadequate/poor image quality or other device-related reasons. The number of injections per patients was similar in the 2 groups (*Control group* 33±24 versus *CVR group* 37±25; p = 0.072). Seven participants (6 in the *Control group* and one in the *CVR group*) did not have at least one in-hospital post-procedural sCr value because they died within 24 (3-22) hours after enrollment (Table S8). All the other participants had at least two post-procedural sCr values (that is at 24 and 48 hours), and 3 o more values were available for 539 (98%) participants. (Figure 2, panel B; Figure S1). AKI occurred in 65/268 (24.3%) participants in the *Control group* and in 44/275 (16%) in the *CVR group* (RR = 0.66; 95% CI 0.47-0.93; p = 0.018). The absolute risk difference was −8.3% (−0.14%, −0.01%). The NNT to prevent one event with the DyeVert system was 12. The association between of CM volume and CA-AKI was also confirmed at graphical analysis (Figure S2).

**Figure 2.**
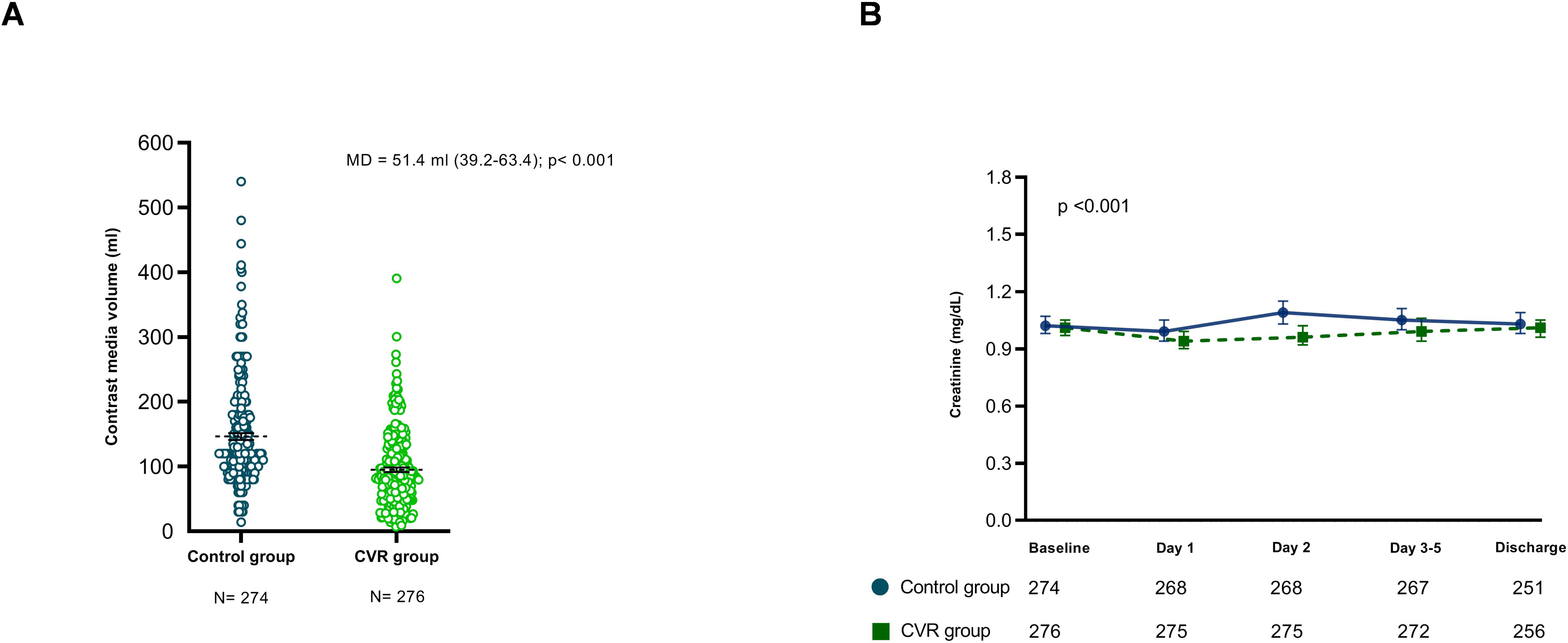
**Panel A**: Cumulative distribution of contrast media volume by group. Black dotted line = mean; Error bars = standard error of means. Difference in means (MD) with 95% CI is reported on top. **Panel B.** Serum creatinine concentrations (geometric means with their 95% confidence intervals) at baseline, day 1, day 2, and days 3-5 after contrast media administration in the *Control group* (dashed line and closed circle) and in the *CVR group* (continuous line and closed square).

### Sensitivity analysis and secondary endpoints

When considering all enrolled participants, and including the 6 participants in the *Control group* who died within 24 hours without AKI, whereas the only participant in the *CVR group* who died within 24 hours with event, AKI occurred in 65/274 (23.7%) participants in the *Control group* and in 45/276 (16.3%) in the *CVR group* (RR = 0.68; 95% CI 0.49-0.96; p = 0.031). An exploratory analysis appraising the interaction between CA-AKI, CM volume and *CVR group* is reported in the Table S9. The distribution of different cutoffs of sCr and cystatin C increase is represented in Table S10. Kinetic of cystatin C is represented in the Figure S3. Distribution of AKI stage was similar in the 2 groups (Table S11). Renal failure requiring RRT occurred in 14/274 (5.1%) participants in the *Control group* versus 7/276 (2.5%) in the *CVR group* (RR = 0.50; 95% CI = 0.20-1.23; p = 0.13). Length of in-hospital stay was similar in the 2 groups (*Control group* 6.6 ± 4.6 versus *CVR group* 6.7 ± 6.1 days; p = 0.78). On the contrary, length of in-hospital stay was longer in participants who experienced AKI (11.1 ± 9.86 vs. 5.6 ± 2.7 days; p<0.001).

The 1-month and 6-month major adverse cardiac and kidney event rate are reported in the Table 3. Sustained kidney damage was higher in the *Control group* than in the *CVR group*.

**Table 3.** Major adverse cardiac and kidney events at 1-month and 6-month in the 2 groups.

|  | **Control group**<br>**(n= 274)** | **CVR group**<br>**(n= 276)** | **P** |
| --- | --- | --- | --- |
| **1-month** |  |  |  |
| **Cumulative major adverse events** | 83 (30%) | 65 (23.5%) | 0.077 |
| **Death** | 23 (8.4%) | 20 (7.2%) | 0.63 |
| **Myocardial infarction** | 3 (1.1%) | 7 (2.5%) | 0.22 |
| **Dialysis** | 14 (5.1%) | 7 (2.5%) | 0.12 |
| **Sustained kidney damage\*** | 67 (24.53%) | 46 (16.73%) | 0.026 |
| **Major bleeding (BARC 3/5)** | 6 (2.2%) | 5 (1.8%) | 0.77 |
| **6-month** |  |  |  |
| **Cumulative major adverse events** | 91 (33%) | 72 (26%) | 0.076 |
| **Death** | 29 (10.5%) | 25 (9.0%) | 0.57 |
| **Myocardial infarction** | 5 (1.8%) | 9 (3.2%) | 0.42 |
| **Dialysis** | 14 (5.1%) | 7 (2.5%) | 0.12 |
| **Sustained kidney damage#** | 69 (25.1%) | 48 (17.4%) | 0.028 |
| **Major bleeding (BARC 3/5)** | 8 (2.9%) | 5 (1.8%) | 0.41 |

\* Median time from contrast media administration to the serum creatinine measurement used to assess sustained kidney damage at 1 month was 30 (IQR 26-30) days in *Control group* (n = 268) and 30 (28-30) days in *CVR group* (n = 275) (p = 0.48).

## DISCUSSION

The pathogenesis of AKI in the setting of ACS is multifactorial. Age, unstable hemodynamic conditions, co-morbidities, pre-existing chronic kidney disease, dehydration, administration of nephrotoxic drugs and CM may concur in the development of AKI ^4^. Recently, the role of iodinated CM has been questioned because the studies linking CM use to AKI in the setting of ACS lack of a control group in which CM was not administered, making thus impossible to distinguish CM-associated AKI from CM-independent AKI. Caspi et al. ^27^ reported that AKI rate was similar in STEMI patients with and without CM exposure. However, this finding comes from an observational nonrandomized study, in which the use of propensity score matching cannot fully correct for imbalances and selection bias. At present, it is unethical and unrealistic to design a randomized, controlled study to assess AKI rate in ACS patients treated by invasive versus medical approach. A way for overcoming this issue is to test whether strategies for CM minimization are associated with a reduction in AKI rate. Indeed both a non-linear and a linear relationship between CM volume and AKI has been reported ^23, 28, 29^. Gurm et al. suggested that a 30% reduction in CM volume could translate into a 12.8% reduction in AKI ^28^. The ultra-low or zero contrast procedures have been advocated and several CM-sparing strategies have been proposed ^22^. All these strategies however are “operator-dependent”. The DyeVert^TM^ system is an additional, “operator-independent” tool contributing to the CM-sparing approach. Use of the DyeVert™ system resulted in a 41% reduction in CM volume^7, 15^

CM volume administered during invasive procedure in ACS patients is on average 138-425 mL^1–3, 5, 18^. In a pooled analysis from the HORIZONS-AMI and the ACUITY trials the CM volume was 232 mL in the CA-AKI group and 220 mL in the group without CA-AKI ^1^. In a STEMI population, Marenzi et al. reported a mean CM volume of 216±73 mL in the group without CA-AKI and 425±148 mL in the CA-AKI group ^5^. In the MATRIX trial mean CM volume was 183±104 mL in the radial access group and 183±110 mL in the femoral access group ^2^. In the present study, CM volume administered in the *Control group was* 160 ± 23 mL, which is one of the lowest values reported in this setting. This finding supports the fact that strategies for CM minimization have been implemented. Moreover, CM volume administered in the *CVR group* was 95 ± 30 mL. The current REMEDIAL IV therefore supports the observation that, on top of all strategies recommended for CM minimization, the DyeVert^TM^ system is effective in limiting CM volume even in ACS patients. This CM volume saving was associated with a significant reduction in CA-AKI rate (absolute risk difference was −8.3%) . This finding 1) supports the concept that CM volume is an important determinant of AKI in ACS patients, and 2) emphasizes the need to implement all strategies effective for CM minimization including the DyeVert^TM^ system.

Volume expansion represents a widely accepted prophylactic strategy for CA-AKI . At present there is no consensus on how volume expansion should be carried out, especially in ACS patients. The most recommended regimen is normal saline infusion at 1 mL/kg/h (0.5 mL/kg/h if LVEF ≤35% or NYHA >2) from 12 hours before to 24 hours after CM exposure ^6^. This regimen, however, is not suitable in urgent/emergent settings. Maioli et al. suggested an early rapid hydration (3 mL/kg/h starting in the emergency room) followed by infusion of 1 mL/kg/h for 12 hours ^18^. However, this forced volume expansion regimen is contraindicated in ACS patients with unstable hemodynamic conditions. The concept of tailored-volume expansion strategy has been increasingly accepted.. In the present trial we adopted the LVEDP-guided protocol ^17^ because this is simple and easy to implement in ACS patients undergoing urgent/immediate invasive approach, as also recently demonstrated in the ATTEMPT trial_30._

### Study limitations

The lack of blinding and the relatively small participants centers may have influenced decisions on number of injections and amount of contrast media volume used. Indeed, the physicians who did the procedure was not masked. However, the laboratory personnel processing the samples and the components of the CEC were blind of the treatment group. Contrast injection was handled by manual syringe in the majority of participants. Automatic injection was used in ≈7% of participants. Therefore, although the CM saved was similar irrespective to the injection strategy, we should be cautious about extending these results to participants in which automatic injection is used.

### Conclusions

The current study suggests that in ACS patients requiring urgent or immediate invasive approach 1) CM volume is an important determinant of AKI and 2) CM minimization obtained by the DyeVert^TM^ system is associated with a significant reduction in AKI rate.

Disclosure. None. The company providing the DyeVert^TM^ system (Osprey Medical Inc. Minnetonka, MN, USA) was not involved in the trial design and conduction.

## Data Availability

Data will be available upon appropriate request

## REFERENCES

1. Giacoppo D, Madhavan MV, Baber U, Warren J, Bansilal S, Witzenbichler B, Dangas GD, Kirtane AJ, Xu K, Kornowski R, et al. Impact of Contrast-Induced Acute Kidney Injury After Percutaneous Coronary Intervention on Short- and Long-Term Outcomes: Pooled Analysis From the HORIZONS-AMI and ACUITY Trials. Circ Cardiovasc Interv. 2015;8:e002475.

2. Ando G, Cortese B, Russo F, Rothenbuhler M, Frigoli E, Gargiulo G, Briguori C, Vranckx P, Leonardi S, Guiducci V, et al. Acute Kidney Injury After Radial or Femoral Access for Invasive Acute Coronary Syndrome Management: AKI-MATRIX. J Am Coll Cardiol. 2017;69:2592–2603.

3. Leoncini M, Toso A, Maioli M, Tropeano F, Villani S, Bellandi F. Early high-dose rosuvastatin for contrast-induced nephropathy prevention in acute coronary syndrome: Results from the PRATO-ACS Study (Protective Effect of Rosuvastatin and Antiplatelet Therapy On contrast-induced acute kidney injury and myocardial damage in patients with Acute Coronary Syndrome). J Am Coll Cardiol. 2014;63:71–79.

4. Ronco C, Cicoira M, McCullough PA. Cardiorenal syndrome type 1: pathophysiological crosstalk leading to combined heart and kidney dysfunction in the setting of acutely decompensated heart failure. J Am Coll Cardiol. 2012;60(12):1031–1042.

5. Marenzi G, Assanelli E, Campodonico J, Lauri G, Marana I, De Metrio M, Moltrasio M, Grazi M, Rubino M, Veglia F, et al. Contrast volume during primary percutaneous coronary intervention and subsequent contrast-induced nephropathy and mortality. Ann Intern Med. 2009;150:170–177.

6. Neumann FJ, Sousa-Uva M, Ahlsson A, Alfonso F, Banning AP, Benedetto U, Byrne RA, Collet JP, Falk V, Head SJ, et al. 2018 ESC/EACTS Guidelines on myocardial revascularization. Eur Heart J. 2018;40:87–165.

7. Desch S, Fuernau G, Poss J, Meyer-Saraei R, Saad M, Eitel I, Thiele H, de Waha S. Impact of a novel contrast reduction system on contrast savings in coronary angiography - The DyeVert randomised controlled trial. Int J Cardiol. 2018;257:50–53.

8. Briguori C, Mariano E, D’Agostino A, et al. Contrast media volume control and acute kidney injury in acute coronary syndrome. Rationale and design of the REMEDIAL IV trial. JSCAI 2023; 2:100980.

9. Thygesen K, Alpert JS, Jaffe AS, Chaitman BR, Bax JJ, Morrow DA, White HD. Fourth Universal Definition of Myocardial Infarction. Circulation. 2018;138:e618–e651.

10. Collet JP, Thiele H, Barbato E, Barthelemy O, Bauersachs J, Bhatt DL, Dendale P, Dorobantu M, Edvardsen T, Folliguet T, et al. 2020 ESC Guidelines for the management of acute coronary syndromes in patients presenting without persistent ST-segment elevation. Eur Heart J. 2021;42:1289–1367.

11. Lawton JS, Tamis-Holland JE, Bangalore S, Bates ER, Beckie TM, Bischoff JM, Bittl JA, Cohen MG, DiMaio JM, Don CW, et al. 2021 ACC/AHA/SCAI Guideline for Coronary Artery Revascularization: A Report of the American College of Cardiology/American Heart Association Joint Committee on Clinical Practice Guidelines. J Am Coll Cardiol. 2022;79:e21–e129.

12. Baran DA, Grines CL, Bailey S, Burkhoff D, Hall SA, Henry TD, Hollenberg SM, Kapur NK, O’Neill W, Ornato JP, et al. SCAI clinical expert consensus statement on the classification of cardiogenic shock. Catheter Cardiovasc Interv. 2019;94:29–37.

13. Declaration of Helsinki. Ethical principles for medical research involving human subjects. J Indian Med Assoc. 2009;107:403–405.

14. Anello C, O’Neill RT, Dubey S. Multicentre trials: a US regulatory perspective. Stat Methods Med Res. 2005;14:303–318.

15. Mehran R, Faggioni M, Chandrasekhar J, Angiolillo DJ, Bertolet B, Jobe RL, Al-Joundi B, Brar S, Dangas G, Batchelor W, et al. Effect of a Contrast Modulation System on Contrast Media Use and the Rate of Acute Kidney Injury After Coronary Angiography. JACC Cardiovasc Interv. 2018;11:1601–1610.

16. Briguori C, Golino M, Porchetta N, Scarpelli M, De Micco F, Rubino C, Focaccio A, Signoriello G. Impact of a contrast media volume control device on acute kidney injury rate in patients with acute coronary syndrome. Catheter Cardiovasc Interv. 2021;98:76–84.

17. Brar SS, Aharonian V, Mansukhani P, Moore N, Shen AY, Jorgensen M, Dua A, Short L, Kane K. Haemodynamic-guided fluid administration for the prevention of contrast-induced acute kidney injury: the POSEIDON randomised controlled trial. Lancet. 2014;383:1814–1823.

18. Maioli M, Toso A, Leoncini M, Micheletti C, Bellandi F. Effects of hydration in contrast-induced acute kidney injury after primary angioplasty: a randomized, controlled trial. Circ Cardiovasc Interv. 2011;4:456–462.

19. Delgado C, Baweja M, Crews DC, Eneanya ND, Gadegbeku CA, Inker LA, Mendu ML, Miller WG, Moxey-Mims MM, Roberts GV, et al. A Unifying Approach for GFR Estimation: Recommendations of the NKF-ASN Task Force on Reassessing the Inclusion of Race in Diagnosing Kidney Disease. J Am Soc Nephrol. 2021;32:2994–3015.

20. Mehran R, Owen R, Chiarito M, Baber U, Sartori S, Cao D, Nicolas J, Pivato CA, Nardin M, Krishnan P, et al. A contemporary simple risk score for prediction of contrast-associated acute kidney injury after percutaneous coronary intervention: derivation and validation from an observational registry. Lancet. 2021;398:1974–1983.

21. Gurm HS, Seth M, Kooiman J, Share D. A novel tool for reliable and accurate prediction of renal complications in patients undergoing percutaneous coronary intervention. J Am Coll Cardiol. 2013;61:2242–2248.

22. Almendarez M, Gurm HS, Mariani J, Jr., Montorfano M, Brilakis ES, Mehran R, Azzalini L. Procedural Strategies to Reduce the Incidence of Contrast-Induced Acute Kidney Injury During Percutaneous Coronary Intervention. JACC Cardiovasc Interv. 2019;12:1877–1888.

23. Gurm HS, Dixon SR, Smith DE, Share D, Lalonde T, Greenbaum A, Moscucci M, Registry BMC. Renal function-based contrast dosing to define safe limits of radiographic contrast media in patients undergoing percutaneous coronary interventions. J Am Coll Cardiol. 2011;58:907–914.

24. Levey AS, Eckardt KU, Dorman NM, Christiansen SL, Hoorn EJ, Ingelfinger JR, Inker LA, Levin A, Mehrotra R, Palevsky PM, et al. Nomenclature for kidney function and disease: report of a Kidney Disease: Improving Global Outcomes (KDIGO) Consensus Conference. Kidney Int. 2020;97:1117–1129.

25. Mehran R, Rao SV, Bhatt DL, Gibson CM, Caixeta A, Eikelboom J, Kaul S, Wiviott SD, Menon V, Nikolsky E, et al. Standardized bleeding definitions for cardiovascular clinical trials: a consensus report from the Bleeding Academic Research Consortium. Circulation. 2011;123:2736–2747.

26. Briguori C, Quintavalle C, De Micco F, Visconti G, Di Palma V, Napolitano G, Focaccio A, Condorelli G. Persistent serum creatinine increase following contrast-induced acute kidney injury. Catheter Cardiovasc Interv. 2018;91:1185–1191.

27. Caspi O, Habib M, Cohen Y, Kerner A, Roguin A, Abergel E, Boulos M, Kapeliovich MR, Beyar R, Nikolsky E, et al. Acute Kidney Injury After Primary Angioplasty: Is Contrast-Induced Nephropathy the Culprit? J Am Heart Assoc. 2017;6: e005715.

28. Gurm HS, Seth M, Mehran R, Cannon L, Grines CL, LaLonde T, Briguori C, Blue Cross Blue Shield of Michigan Cardiovascular C. Impact of Contrast Dose Reduction on Incidence of Acute Kidney Injury (AKI) Among Patients Undergoing PCI: A Modeling Study. J Invasive Cardiol. 2016;28:142–146.

29. Narula A, Mehran R, Weisz G, Dangas GD, Yu J, Genereux P, Nikolsky E, Brener SJ, Witzenbichler B, Guagliumi G, et al. Contrast-induced acute kidney injury after primary percutaneous coronary intervention: results from the HORIZONS-AMI substudy. Eur Heart J. 2014;35:1533–1540.

30. Liu Y, Tan N, Huo Y, Chen S, Liu J, Chen YD, Wu K, Wu G, Chen K, Ye J, et al. Hydration for prevention of kidney injury after primary coronary intervention for acute myocardial infarction: a randomised clinical trial. Heart. 2022;108:948–955.

